# SARS-CoV-2 Omicron variant, lineage BA.1, is associated with lower viral load in nasopharyngeal samples compared to Delta variant

**DOI:** 10.1101/2022.02.02.22269653

**Authors:** Célia Sentis, Geneviève Billaud, Antonin Bal, Emilie Frobert, Maude Bouscambert, Gregory Destras, Laurence Josset, Bruno Lina, Florence Morfin, Alexandre Gaymard, the COVID-Diagnosis HCL Study Group

## Abstract

High viral load in upper respiratory tract specimens observed for Delta cases may contributed to its increased infectivity compared to the Alpha variant. Herein, we showed that the RT-PCR Ct values in Health Care Workers sampled within five days after symptom onset were significantly higher for Omicron cases than Delta cases (+2.84 Ct, p=0.008). This result comfort the studies showing that the increased transmissibility of Omicron is related to other mechanisms than higher virus excretion.

---

At the end of 2020, the first SARS-CoV-2 variants of concern (VOC), named Alpha, was detected and became the main variant just a few months after (1,2). Then, Delta variant firstly detected in March 2021 rapidly spread worldwide and became the major variant during the second part of 2021. These VOCs demonstrated increased infectivity that was related to better affinity for ACE2 cellular receptor and higher viral load in respiratory tract samples (3–7).

During the last trimester of 2021, a new variant named Omicron, emerged and was immediately classified as a VOC due to the large number of mutations found in the spike protein, including several mutations known to be associated with higher transmissibility and/or immune escape (8,9). The Omicron variant is the most contagious form of SARS-CoV-2 known so far and became dominant worldwide in a few weeks (10). However, it is not yet documented if its enhanced infectivity is also related to a higher viral load as reported for other variants(5,6).

## Sample collection and virological testing

To determine if the Omicron variant’s spread is related to higher viral loads compared to Delta variant, we collected nasopharyngeal swabs from screening center dedicated to health care workers and family at the University Hospital of Lyon, France. Data from samples taken between 12/1/2021 and 12/31/2021 were collected, a period with circulation of Delta variant and emergence of Omicron variants in Lyon, France (Figure 1A). The French national strategy includes screening test specific for SARS-CoV-2, positive samples are then tested by RT-PCR targeting some mutations (E484K, L452R and K417N) and by whole genome sequencing. SARS-CoV-2 detection was performed using the cobas® 6800 SARS-CoV-2 assay (Roche, Switzerland). Cycle threshold (Ct) for the RdRp target was used as a proxy to evaluate SARS-CoV-2 viral load. Mutations screening were performed using TaqMan SARS-CoV-2 Mutation Panel (Thermofisher, USA) and whole genome sequencing with COVIDSeq assay (Illumina, USA). All statistical analyses were conducted using GraphPad Prism® (version 8.0.2). Mean Ct difference between groups was assessed using the Student’s T test or Mann-Whitney test, as appropriate. Categorical variables were compared using the Chi-square test or Fisher’s exact test.

**Figure 1Aa:**
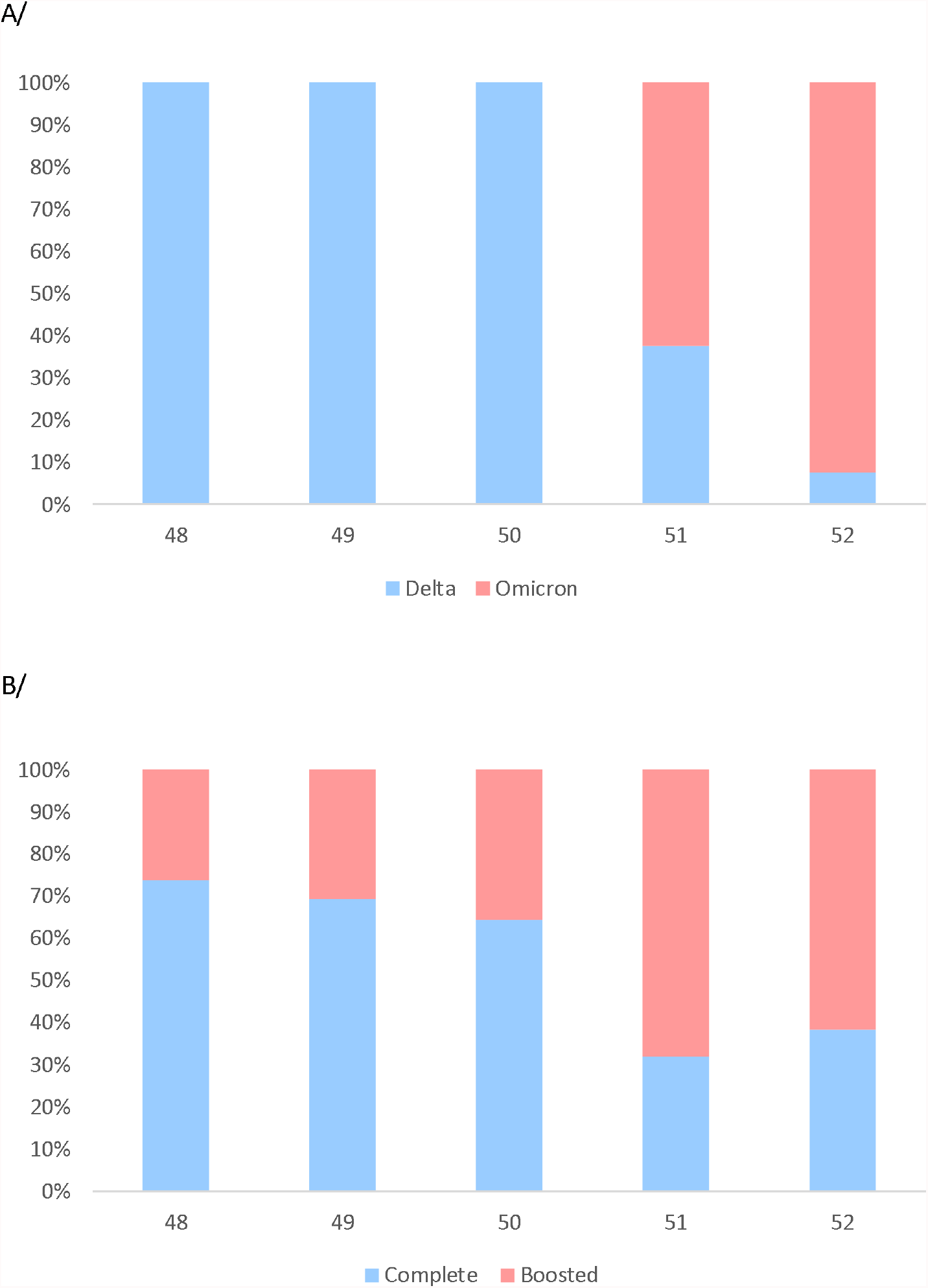
Proportion of samples positive for the Delta or Omicron variant on a weekly basis. Week 48 (Delta, n=31), week 49 (Delta, n=16), week 50 (Delta, n=16); week 51 (Delta, n=18 ; Omicron, n=30), week 52 (Delta, n= 8 ; Omicron, n=99). Figure 1B: Vaccination status of patients on a weekly basis. Patients not vaccinated, partially vaccinated, or with unknown status were removed from this graph. Week 48 (complete, n= 14: boosted, n = 5), week 49 (complete, n =9 ; boosted, n =4), week 50 (complete, n= 9 ; boosted, n =5), week 51 (complete, n = 8 ; boosted, n = 17), week 52 (complete, n = 31 ; boosted, n= 50).

## Ct-values according to day post-symptom onset and age

Patients infected with Omicron variant had a lower viral load (higher Ct value) compared to patients infected with Delta variant (22.7 for Delta vs 24.4 for Omicron, p=0.006) (Figure 2A). This observation was only confirmed for patients with symptoms appearing less than 5 days before sampling (21.7 for Delta vs 23.8 for Omicron, p=0.008). Moreover, the largest difference was found in patients with symptom onset under 1 day, with viral load almost 1 log_10_ lower with Omicron variant compared to Delta variant (21.3 for Delta vs 24.2 for Omicron, p = 0.035) (Figure 2A and Supplementary 1). Significant viral load differences were found only for patients over 40 years old (20.9 for Delta vs 23.6 for Omicron, p=0.006) (Supplementary 1, Figure 2B) with higher viral load found in patients infected by Delta variant.

**Figure 2:**
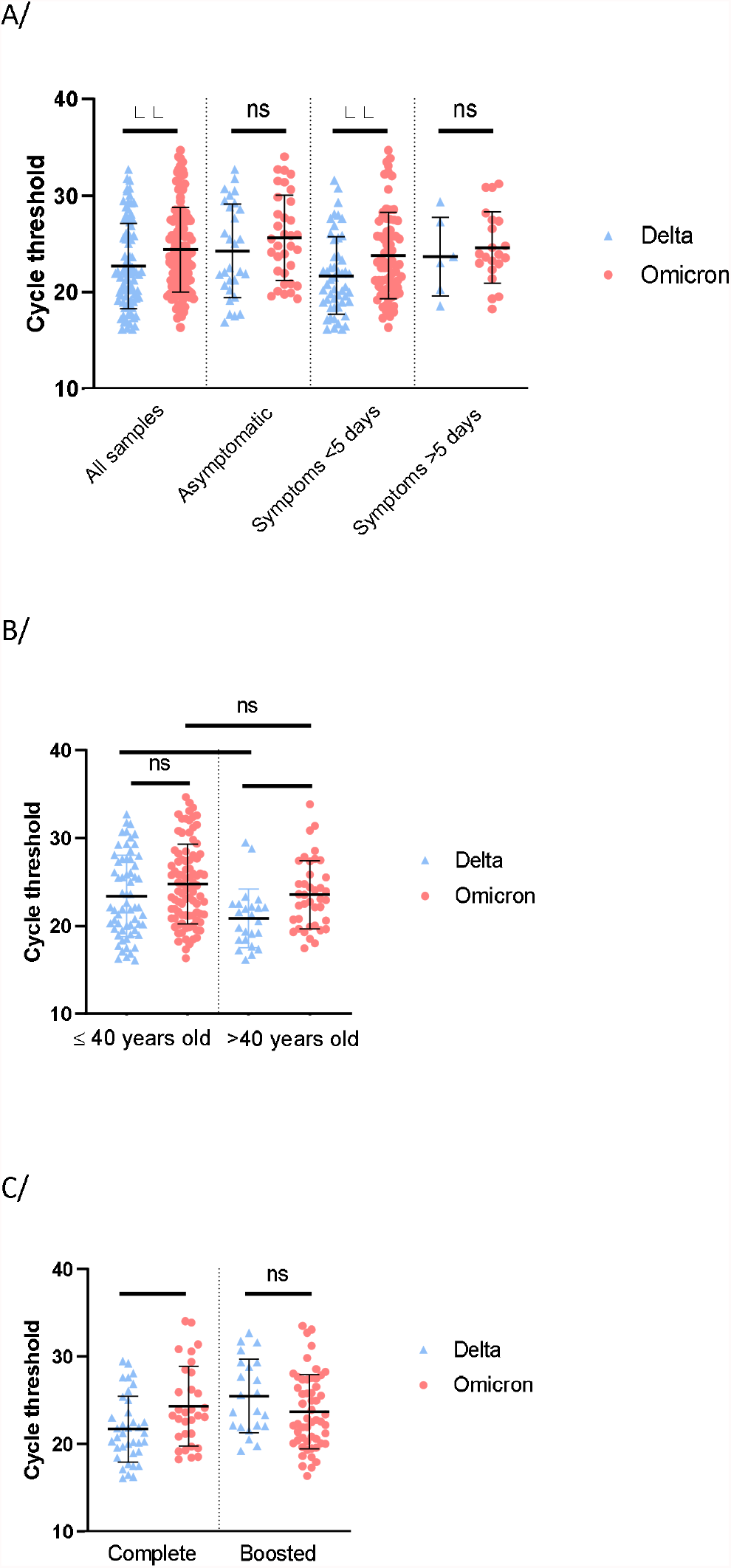
RT-PCR Cycle threshold values for Delta or Omicron 2A: Cycle threshold of Delta or Omicron variant according to symptoms. Global analysis included all samples (Delta, n=86, Omicron, n=129). Asymptomatic patients (Delta, n=29 and Omicron, n=34), symptoms appearing less than 5 days prior sampling (Delta, n=51 and Omicron, n=74) and symptoms appearing more than 5 days prior sampling (Delta, n=6 and Omicron, n=21). 2B: Cycle threshold of Omicron or Delta variant by age. Ct was analysed for patients under 40 years old (Delta, n= 62 and Omicron, n=90) and patients over 40 years old (Delta, n=24 and Omicron, n=39). 2C: Cycle threshold of Delta or Omicron variant according to vaccination status. Vaccination was considered complete when patient received 2 doses or 1 dose and 1 infection (Delta, n= 37 and Omicron, n=31) and boosted when patients received 3 doses or 2 doses and 1 infection (Delta, n=22 and Omicron, n=54). P-value was calculated with Student test. ns = not significant, * = <0.05, ** = <0.01.

## Ct-values according to vaccination status

In order to limit misinterpretation related to the vaccination status, we classified patients into five categories according to French vaccination strategy: not vaccinated, partially vaccinated (1 dose or 1 infection), completely vaccinated (2 doses or 1 dose and 1 infection), boosted (3 doses or 2 doses and 1 infection) or unknown (data not available). Regarding patients completely vaccinated, Omicron variant was characterized by a lower viral load compared to patients infected by the Delta variant (21.7 for Delta vs 24.3 for Omicron, p=0.01). In boosted patients the inverse trend was observed with a higher viral load during Omicron variant infection (25.49 for Delta vs 23.69 for Omicron p=0.09) (Figure 2C). Of note, the proportion of third dose (boosted) was higher at the end of December compared to the beginning of December (26.3% week 48 vs 61.7% week 52, Figure 1B).

## Discussion

For the previous Alpha and Delta VOCs higher transmission rates have been related to higher viral loads (5–7). In contrast, our results showed a higher Ct value (+2.84), reflecting lower viral load (−0.85 log_10_), for patients infected by Omicron, compared to patients infected by Delta variant. This is in agreement with a recent study reporting patients follow-up after Delta and Omicron infection showing a peak viral load at a Ct value of 23.3 for Omicron and 20.5 for Delta (11). In another study, Puhach *et al*, reported a low correlation between RNA genome copies and infectious virus shedding evaluated by viral culture (12). Regarding Omicron, they showed a trend to lower viral load compared to Delta (RNA genome copies and infectious virus titers) that was not significant probably due to low patients number in the Omicron group (n=18) (12). These data combined with ours could suggest that higher infectiousness of Omicron may not be related to an increased viral load as reported for previous variants. Complete mechanisms driving the higher transmissibility of Omicron variant are still unknown. Infectiousness could be multifactorial and related to background immunity, respiratory symptoms (cough, sneeze), duration of viral excretion, age and viral parameters such as new viral entry mechanism (11,13,14).

The present study has several limitations as viral load was estimated by Ct without specific quantification and normalization. Only BA.1 lineage were circulating in France during the study period and results for other Omicron lineage such as BA.2 could differ. Our results were also impacted by the vaccination strategy. Omicron viral load was lower for patients with complete vaccination but the inverse trend was observed for boosted patients. This might be related to a lower susceptibility to neutralizing antibodies for Omicron variant compared to Delta variant. Boosted patients infected by Delta variant might be more protected than patients infected by Omicron, which would explain the lower viral load in Delta group (8,15,16). These results should be taken carefully as more patients with boosted vaccination were observed in the Omicron infection group and counterwise more patients with complete vaccination were observed in the Delta infection group (Table 1). This observation has to be related to the National vaccination strategy, as French government announced a mandatory third dose vaccination to health care workers during December. In addition, most of positive samples for Delta variant were collected at the beginning of December. Omicron variant began to be detected in France mid-December 2021 and overthrow Delta variant during the last week of December (Figure 1A). A more in-depth study taking into account vaccination status, time since the last injection and antibody levels will be necessary to better understand the impact of the immune response on the infection by different SARS-CoV-2 variants.

**Table 1:** Demographic data. P-value was calculated with Fischer test.

|  | Delta<br>(n=86) | Omicron<br>(n=129) | P-value |
| --- | --- | --- | --- |
| Sex |  |  |  |
| Women | 65.1 % (56) | 62.8% (81) | 0.77 |
| Men | 34.9% (30) | 37.2% (48) |  |
| Index ratio | 1.86 | 1.68 |  |
| Age |  |  |  |
| 20-30 | 38.4% (33) | 45.8% (59) | 0.32 |
| 31-40 | 33.7% (29) | 24.0% (31) | 0.12 |
| 41-50 | 16.3% (14) | 18.6% (24) | 0.71 |
| > 51 years old | 11.6% (10) | 11.6% (15) | > 0.99 |
| Symptoms |  |  |  |
| Asymptomatic | 33.7% (29) | 26.3% (34) | 0.28 |
| Day before or day of sampling | 24.4% (21) | 28.7% (37) | 0.53 |
| 2 to 4 days before sampling | 34.9% (30) | 28.7% (37) | 0.37 |
| 5 to 7 days before sampling | 5.8% (5) | 12.4% (16) | 0.16 |
| 8 to 14 days before sampling | 1.2% (1) | 3.9% (5) | 0.4 |
| Vaccination status |  |  |  |
| Not vaccinated | 2.3% (2) | 5.4% (7) | 0.32 |
| Partial : 1 dose or 1 infection | 8.1% (7) | 6.9% (9) | 0.79 |
| Complete : 2 doses or 1 dose + 1 infection | 43% (37) | 24.0% (31) | **0.004** |
| Boosted : 3 doses or 2 doses + 1 infection | 25.6% (22) | 41.9% (54) | **0.02** |
| Unknown | 20.9% (18) | 21.7% (28) | > 0.99 |

Finally, lower viral load during Omicron infection might impact viral diagnosis. Even if rapid antigenic testing are still able to detect Omicron, higher Ct values in Omicron cases may be associated with an increased number of false negative results compared to Delta. This must not preclude from using antigenic testing devices, but the interpretation may be cautious. Moreover, patient monitoring using Ct values should be cautiously interpreted according to each patient situation. Ct value could be a poor indicator of infectiousness, especially in presence of neutralizing antibodies. In the context of a largely vaccinated population, new criteria must be defined and new biomarkers have to be looked for.

## Data Availability

All data produced in the present study are available upon reasonable request to the authors

## Conflic of interest

None

## Funding statement

No specific funding was obtain for this study.

## Data availability

SARS-CoV-2 whole genomes sequenced in this study were deposited in the GISAID database.

## Ethics statement

Ethics committee of Hospices Civils de Lyon gave ethical approval for this work (approval number 22_787). The investigations were carried out in accordance with the General Data Protection Regulation (Regulation (EU) 2016/679 and Directive 95/46/EC) and the French data protection law (Law 78–17 on 06/01/1978 and Décret 2019–536 on 29/05/2019). Samples were collected for regular clinical management, with no additional samples for the purpose of this study. Patients were informed and their non-objection approval was confirmed.

## COVID-Diagnosis HCL Study Group

Jean-Sébastien Casalegno, Vanessa Escuret, Vinca Icard, Marion Jeannoel, Marie-Paule Milon, Yahia Mekki, Christophe Ramière, Caroline Scholtès, Bruno Simon, Jean-Claude Tardy, Mary-Anne Trabaud, Isabelle Schuffenecker, Martine Valette.

All authors in the study group are affiliated to :

Laboratoire de Virologie, Institut des Agents Infectieux, Laboratoire associé au Centre National de Référence des virus des infections respiratoires, Hospices Civils de Lyon, Lyon,

## Authors’ contributions

CS, FM, and AG designed the study. GB, EF, MB supervised the RT-PCR analysis. AB GD LJ and AG supervised the variant identification. AG wrote the protocol of the study. CS, FM and AG collected the data. CS, BL, FM and AG analysed the whole results. CS performed the statistical analyses and provided the Figures and Tables. The members of the COVID-Diagnosis HCL Study Group contributed to the laboratory analysis. CS, BL, FM and AG wrote the manuscript. All the authors corrected and approved the final manuscript.

## Notes

### Competing Interest Statement

The authors have declared no competing interest.

### Funding Statement

This study did not receive any funding

### Author Declarations

Ethics committee of Hospices Civils de Lyon gave ethical approval for this work (approval number 22_787). The investigations were carried out in accordance with the General Data Protection Regulation (Regulation (EU) 2016/679 and Directive 95/46/EC) and the French data protection law (Law 78.17 on 06/01/1978 and Decret 2019.536 on 29/05/2019). Samples were collected for regular clinical management, with no additional samples for the purpose of this study. Patients were informed and their non-objection approval was confirmed.

