## Supplementary table 1 for "SARS-CoV-2 Omicron variant, lineage BA.1, is associated with lower viral load in nasopharyngeal samples compared to Delta variant"

|  | **Delta** | | **Omicron** | |  |
| --- | --- | --- | --- | --- | --- |
| **RT-PCR SARS-CoV-2 COBAS** | **Ct** | **S_D_** | **Ct** | **S_D_** | **p-value** |
| All samples | 22.70 | 4.44 | 24.4 | 4.38 | **0.006** |
| **Symptoms** |  |  |  |  |  |
| Asymptomatic prior samples | 24.27 | 4.85 | 25.62 | 4.42 | 0.25 |
| Symptoms <5 days prior samples | 21.7 | 4.02 | 23.79 | 4.47 | **0.008** |
| Symptoms day before or day of sample | 21.35 | 3.85 | 24.19 | 5.31 | **0.035** |
| Symptoms 2 to 4 days before sample | 21.94 | 4.18 | 23.38 | 3.46 | 0.13 |
| Symptoms >5 days prior sample | 23.69 | 4.07 | 24.61 | 3.71 | 0.6 |
| **Vaccination** |  |  |  |  |  |
| Complete vaccination scheme (2 doses or 1 dose + 1 infection) | 21.72 | 3.77 | 24.33 | 4.58 | **0.01** |
| Boosted vaccination schema (3 doses or 2 doses + 1 infection) | 25.49 | 4.22 | 23.69 | 4.25 | 0.09 |
| **Sex** |  |  |  |  |  |
| Women | 22.61 | 4.48 | 24.37 | 4.22 | **0.02** |
| Men | 22.88 | 4.43 | 24.46 | 4.69 | 0.14 |
| **Age** |  |  |  |  |  |
| < 30 years old | 23.69 | 4.57 | 24.77 | 4.67 | 0.28 |
| 31-40 years old | 23.11 | 4.75 | 24.77 | 4.38 | 0.16 |
| 41 -50 years old | 20.53 | 2.28 | 23.32 | 3.64 | **0.01** |
| > 51 years old | 21.34 | 4.53 | 23.93 | 4.35 | 0.16 |
| ≤40 years old | 23.41 | 4.63 | 24.77 | 4.55 | 0.07 |
| >40 years old | 20.87 | 3.34 | 23.56 | 3.88 | **0.006** |

Supplementary 1 : Cycle threshold analysis between Delta and Omicron variant according to symptoms, vaccination status, sex and age. P-value was calculated with a Student test.
